# Impact of exposures to persistent endocrine disrupting compounds on the sperm methylome in regions associated with neurodevelopmental disorders

**DOI:** 10.1101/2021.02.21.21252162

**Authors:** Angela G. Maggio, Henry T. Shu, Benjamin I. Laufer, Hyeyeon Hwang, Chongfeng Bi, Yinglei Lai, Janine M. LaSalle, Valerie W. Hu

## Abstract

**Background:** Although autism spectrum disorder (ASD) is among the most heritable of neurodevelopmental disorders, the rapidly rising prevalence of ASD suggests that environmental factors may interact with genetic risk for ASD. Environmental factors may impact both gene expression and phenotypes in ASD through epigenetic modifications that, in turn, could lead to intergenerational effects influencing risk for ASDs. Endocrine disrupting compounds (EDCs), such as the long-lived organochlorines, are of particular interest with respect to risk for autism because of their ability to interfere with sex hormones that have been implicated in the regulation of *RORA*, a dysregulated gene in ASD that is a master regulator of many other ASD risk genes.

**Objectives:** The specific aims of this study are to: 1) investigate whether high versus low exposures to the persistent organochlorine 1,1-dichloro-2,2-bis(p-chlorophenyl)ethylene (DDE) are associated with differentially methylated regions (DMRs) in sperm from a Faroese cohort whose natural diet of pilot whale meat and blubber exposes them to higher than average levels of organic pollutants; 2) determine if genes associated with DDE DMRs are enriched for ASD risk genes; 3) identify pathways and functions over-represented among genes associated with DMRs.

**Methods:** Whole genome bisulfite sequencing (WGBS) was used to identify genome-wide DMRs in sperm from individuals divided by high and low exposure levels. Gene ontology and pathway analyses were used to determine enrichment in functional relationships to ASD.

**Results:** Genes in DMRs not only could discriminate between high and low exposures to DDE, but also were enriched in autism risk genes. Gene ontology and pathway analyses of these genes show significant enrichment for neurodevelopmental processes frequently impacted by ASD.

**Conclusion:** Results of this study show that elevated exposure to certain organochlorines is associated with genome-wide DNA methylation patterns in sperm affecting genes involved in neurological functions and developmental disorders, including ASD.

## Introduction

Autism spectrum disorder (ASD) is a highly complex neurodevelopmental disorder characterized by deficits in social communication, repetitive behaviors, and restricted interests (American Psychiatric Association. Task Force on DSM-5, 2013). In a span of only 12 years from 2004 to 2016, the United States’ Center for Disease Control and Prevention reported an increase in prevalence of ASD from roughly 1 in 125 children to 1 in 54, with a male to female sex bias of 4.3 to 1 (Maenner et al., 2020). While increased awareness of the disorder and expansion of the diagnosis to include the moderate and milder forms of ASD (such as pervasive developmental disorder-not otherwise specified and Asperger syndrome) has been suggested to account for the increase in prevalence, epidemiological studies suggest that there may be environmental contributions to ASD as well (Nevison, 2014). Similarly, although ASD exhibits a strong genetic component as revealed by concordance rates in studies on affected monozygotic versus dizygotic twins (Hallmayer et al., 2011; Tordjman et al., 2014), the incomplete penetrance of ASD even in monozygotic twins also suggests a role for gene by environment (G x E) interactions which are often mediated through epigenetic mechanisms.

Early evidence for the involvement of epigenetics in neurodevelopmental disorders comes from studies on syndromic forms of ASD (e.g., Fragile X, Angelman’s, Prader-Willi, and Rett syndromes) in which the etiologically relevant genes are either aberrantly methylated/imprinted or are involved in the recognition of methylated DNA sequences (Grafodatskaya, Chung, Szatmari, & Weksberg, 2010; Rangasamy, D’Mello, & Narayanan, 2013). In 2010, we demonstrated for the first time genome-wide DNA methylation differences associated with idiopathic autism in lymphoblastoid cell lines (LCLs) from monozygotic twins and sib pairs discordant for diagnosis of autism (Nguyen, Rauch, Pfeifer, & Hu, 2010). Importantly, by integrating these large-scale methylation analyses with genome-wide transcriptome analyses of the same cohort, we showed that the majority of the methylation differences were inversely associated with differences in gene expression (Hu, Frank, Heine, Lee, & Quackenbush, 2006; Hu et al., 2009). Moreover, two of the validated hypermethylated genes, *BCL2* and *RORA*, were also found to be reduced in brain tissues from individuals with ASD (Nguyen et al., 2010), thus linking these molecular changes observed in blood-derived cells to autism brain pathology. We further showed that *RORA* could be regulated in opposite directions by male and female sex hormones, with male hormones suppressing expression (Sarachana, Xu, Wu, & Hu, 2011). In addition, we demonstrated that over 2000 genes were putative transcriptional targets of this nuclear hormone receptor, including hundreds of autism risk genes (Sarachana & Hu, 2013). These findings led us to postulate that this hormonal dependence of *RORA* expression might contribute to the observed sex bias in ASD that has been hypothesized to be related to elevated fetal testosterone or steroidogenic activity (Baron-Cohen et al., 2014; Simon Baron-Cohen, Knickmeyer, & Belmonte, 2005). Moreover, we postulated that endocrine disrupting compounds (EDCs) that can mimic or interfere with normal hormonal signaling and metabolism might perturb the expression and function of this critical candidate gene for ASD, thereby increasing risk for autism (Hu, 2012).

Indeed, environmental exposures to EDCs have been linked with various diseases including neurodevelopmental disorders (Braun, 2017; Gore, Martien, Gagnidze, & Pfaff, 2014; Rivollier, Krebs, & Kebir, 2019; Schug, Blawas, Gray, Heindel, & Lawler, 2015). Both experimental and epidemiological studies have reported harmful effects of exposures to both short- and long-lived EDCs, such as bisphenol A (BPA) and phthalates, and persistent organic pollutants (POPs) like polychlorinated biphenyl (PCB) and polybrominated diphenyl ether (PBDE) compounds, on neurodevelopment. Rodent studies have shown associations between EDC exposure and disruption of social hormones, social recognition, locomotion, and excitatory-inhibitory synapse pathways (Gogolla et al., 2009; Lichtensteiger et al., 2015; Wolstenholme, Goldsby, & Rissman, 2013). Exposure to 3,3’-dichlorobiphenyl exerted effects on axonal and dendritic growth in primary rat neurons (Sethi et al., 2017). Epidemiological studies further show that prenatal human exposures to EDCs, specifically DDE and PCBs, has been linked to alterations in hormone levels in offspring with potential for lasting and widespread signaling disruption (Braun, 2017; Eskenazi et al., 2017).

With respect to mechanisms of action, many studies have investigated epigenetics as a potential mediator of EDC-induced changes in phenotype and disease state. For example, Manikkam et al. showed that certain plastics-derived EDCs induced transgenerational inheritance of obesity, reproductive disease and epimutations in sperm (Manikkam, Guerrero-Bosagna, Tracey, Haque, & Skinner, 2012). More recently, McBirney et al. reported that exposure of pregnant rats during gestation to the herbicide atrazine increased the frequency of testis disease and mammary tumors as well as induced changes in body weight and onset of puberty in the F2 and F3 offspring (McBirney et al., 2017). Additionally, they reported DNA methylation changes in sperm, which were detected in offspring in each generation (McBirney et al., 2017). Regarding EDC involvement in autism, Dunaway et al. demonstrated that treatment of a neuronal cell model with PCB 95 was associated with significant global DNA hypomethylation of autism candidate genes with a direct impact on gene expression (Dunaway et al., 2016). In the same study, hypomethylation was also observed in brain tissues from individuals with Dup15q syndrome (a condition frequently associated with ASD) which had been previously associated with exposure to PCB 95 (Mitchell et al., 2012). The role of epigenetics as an underlying mechanism for EDC-induced neurodevelopmental disorders and ASD is further discussed in a number of recent reviews (Bakulski, Singer, & Fallin, 2014; Lasalle, 2013; Moosa, Shu, Sarachana, & Hu, 2018; Tran & Miyake, 2017).

In addition to having an impact on neurodevelopment, EDCs have also been implicated in many other diseases or human conditions, such as cancer, diabetes, obesity, metabolic disorders, and infertility (De Coster & Van Larebeke, 2012). With respect to infertility, human exposures to a variety of organochlorines have been shown to affect chromatin integrity (Rignell-Hydbom et al., 2005) and sex chromosome aneuploidy in sperm (McAuliffe, Williams, Korrick, Altshul, & Perry, 2012). EDC-associated sex chromosome aneuploidy in sperm was further confirmed in an additional study involving a cohort of men in the Faroe Islands who have higher than average exposures to POPs based on their diet which is rich in pilot whale meat and blubber (Perry et al., 2016). These chromosomal changes in sperm suggested that EDCs may also induce epigenetic changes in sperm DNA.

In recent years, the sperm methylome (i.e., genome-wide DNA methylation pattern) has come under increasing scrutiny as a reservoir of epigenetic changes that may have an impact on a wide variety of human disorders, from infertility to neurodevelopmental and psychiatric disorders (Wu, Hauser, Krawetz, & Pilsner, 2015). In particular, DNA methylation changes in sperm have been reported as a function of advanced paternal age, in both mice and men (Jenkins, Aston, Pflueger, Cairns, & Carrell, 2014; Milekic et al., 2014). Notably, the changes have included genes that overlap with those reportedly associated with autism, schizophrenia, and bipolar disorder. Since advanced paternal age has also been associated with risk for autism (Quinlan, McVeigh, Driver, Govind, & Karpati, 2015; Reichenberg et al., 2006; Vierck & Silverman, 2014), Feinberg et al. recently investigated DNA methylation differences in sperm from fathers with a child with ASD and from fathers of neurotypical children (Feinberg et al., 2015). They identified 193 DMRs in paternal sperm of ASD fathers, many of which were in the proximity of genes involved in developmental processes. A more recent study on the sperm methylome of fathers that have children with or without ASD further shows the potential for development of a biomarker screen for ASD based on DMRs (Garrido et al., 2021). These studies thus establish a link between DNA methylation changes in sperm and possible risk for ASD. However, it does not address potential environmental contributors to such methylation changes in sperm of the fathers with children affected by ASD.

The primary objectives of this study are to: 1) determine if high versus low exposures to the persistent organochlorine 1,1-dichloro-2,2-bis(p-chlorophenyl)ethylene (DDE) are associated with differentially methylated regions (DMRs) in sperm from a Faroese cohort; 2) determine if genes associated with DDE DMRs are enriched for ASD risk genes; 3) identify pathways and functions among genes associated with DMRs.

## Materials and methods

### Semen samples and demographics of donors

Raw ejaculate (i.e., semen) from 52 men in a general population cohort of the Faroe Islands (Denmark) was kindly provided by Dr. Pál Weihe, Director of the Department of Occupational Medicine and Public Health of the Faroese Hospital System in Tórshavn, Faroe Islands. Inhabitants of the Faroe Islands are considered genetic isolates and therefore are expected to have similar genetic background and polymorphisms that would otherwise be considered major confounding variables in genomics studies. Samples were collected throughout 2007 and 2008 and de-identified by the Faroese Hospital System. Information related to height, weight, age, smoking status, length of abstinence, sample collection date, serum concentrations of the four most prevalent PCB congeners, (PCB 118, PCB 138, PCB 153, PCB 180) and DDE, the major breakdown product of the insecticide DDT (dichloro-diphenyl-trichloroethane) were recorded for each sample as part of the Faroese General Population research cohort (which was part of the parent study approved by the local Science Ethics Committee for the Faroe Islands). The de-identified semen samples were made available for this study through a Data Processor Agreement between Dr. Pál Weihe at the Faroese Hospital System and Dr. Valerie Hu at The George Washington University, together with the serum levels of the organochlorines that had been determined for each donor as previously described (Grandjean et al., 2001). A summary of demographic information and serum concentrations of EDCs for the cohort used in this study is provided in **Supplemental Table 1**. Regression analyses of the potential covariates were performed using statistical packages in Excel. These analyses showed that while there were significant correlations between the concentrations of DDE with those of DDT and the sum of the four most prevalent PCB congeners (**Supplemental Figure 1**), there were no correlations between DDE levels and body mass index (BMI), smoking status, or any of the sperm parameters, including sperm concentration and mobility (**Supplemental Figures 2 and 3**). Faroese semen samples with a minimum sperm count of 20 x 10^6^ per ml were divided by Dr. Melissa Perry (GWU) into three exposure groups based on the recorded levels of DDE in serum, and 26 samples each from the first (lowest exposure) and third (highest exposure) tertiles were provided for this methylation study. It should be noted that, once the 52 semen samples were received, they were processed through bisulfite sequencing analyses without knowledge of the exposure tertiles to which they were assigned.

### Sperm isolation and DNA extraction methods

Sperm was isolated from semen (which contains various cell types) using a published discontinuous gradient protocol (Wu, de Gannes, Luchetti, & Richard Pilsner, 2015). Briefly, 100 μl of semen was washed with 2 mL Quinn’s Sperm Washing Medium (Origio, Trumbull, CT), and cells were pelleted at 600 g for 5 minutes at 4°C. The pelleted cells were resuspended in 0.5 ml Quinn’s solution and counted in a hemocytometer to determine the initial number of sperm and somatic cells in the semen sample. Next, a discontinuous gradient of PureCeption solution (Origio) diluted with Quinn’s was formed in a 15 ml conical tube with 1.5 ml of 40% PureCeption over 1.5 ml of 90% PureCeption. The washed cells were layered on top of the gradient before centrifugation at 300 g for 20 minutes at room temperature. The pellet was transferred to a new 15 ml tube and resuspended in 3 ml Quinn’s Washing solution. The cells were pelleted at 600 g for 5 minutes, and then resuspended in 500 μl Quinn’s after removal of the supernatant. Cells were recounted to determine total number of sperm and somatic cell contamination (which never exceeded 1%) before centrifugation at 4000 g for 1 minute.

DNA was isolated from the purified sperm cells using a Qiagen AllPrep DNA/RNA mini kit following the manufacturer’s protocol. Sperm pellets were first lysed in 450 μl RLT buffer with 50 μl added tris(2-carboxyethyl)phosphine solution (TCEP, a bond-breaker) by vortexing with 0.1 gm RNase/DNase-free stainless steel microbeads for 5 min at RT in a Disruptor Genie (Scientific Industries, Bohemia, NY). Lysates were immediately stored at −80°C until DNA extraction which was usually completed the next day using the standard protocol.

### Whole Genome Bisulfite Sequencing (WGBS)

Samples for WGBS were divided into two batches, a discovery set of 32 samples and a validation set of 20 samples. The samples were sent to Admera Health (South Plainfield, NJ), a CLIA-certified laboratory, for WGBS analyses. The directional Illumina TruSeq DNA Methylation library kit was used for sample preparation. WGBS was then performed on an Illumina HiSeq X sequencer resulting in 150 bp PE reads achieving roughly 4x coverage genome wide per sample. Raw FASTQ files were received from Admera Health for further analyses. As mentioned previously, all samples were processed blindly without the processors’ knowledge of tertile level from sperm isolation through the WGBS analyses to minimize handling bias.

### WGBS bioinformatics pipeline

We utilized a bioinformatics pipeline comprised of CpG_Me for alignment and DMRichR for differential methylation determination as published on github (https://github.com/ben-laufer). CpG_Me builds on previously published bioinformatic tools and pipelines (Krueger & Andrews, 2011; Laufer, Hwang, Vogel Ciernia, Mordaunt, & LaSalle, 2019). The DMRichR workflow similarly builds on previously established bioinformatic packages such as dmrseq and bsseq (Hansen, Langmead, & Irizarry, 2012; Korthauer, Chakraborty, Benjamini, & Irizarry, 2019; Laufer et al., 2019). All WGBS data was analyzed on The George Washington University’s high-performance cluster, Colonial One. First, raw read files (FASTA) were trimmed and quality checked with Trim-Galore and FASTQC software. Forward reads were trimmed by 8bp on the 3’- and 5’-ends. Reverse reads were trimmed 8bp on the, 3’-ends and 20bp on the 5’-ends in order to remove methylation bias often seen at the ends of reads. M-Bias plots from FASTQC analyses were examined to determine if trimming was sufficient. Trim Galore was also used to filter out bases with Phred scores lower than 20 that would indicate a 1 in 100 probability of an incorrect base call. Reads were then aligned to a reference human genome (hg38) using Bismark, a three-letter aligner. Methylation calls are differentiated among CpG, CHG, and CHH contexts (Krueger & Andrews, 2011), but only the CpG sites were considered in this study. Approximately 74% of bisulfite reads aligned to a bisulfite-converted reference human genome allowing for the assay of 10.08 million CpG sites. All raw and processed data from the WGBS analyses have been deposited into the NBCI’s Gene Expression Omnibus (GEO) repository (GEO Accession number GSE165915).

### Identification of differentially methylated regions (DMRs)

DMRs were identified using dmrseq and bsseq Bioconductor packages in the wrapped pipeline of DMRichR. Default parameters for the DMRichR executable script were used which included coverage set to 1x, per Group set to 100%, minCpGs set to 5, and maxPerms set to 10. Covariables adjusted for in DMR analysis included BMI, age, days in storage, batch effects of processor and date of processing, percent motile sperm, and smoking status. With DMRichR, DMRs were annotated for genomic and CG context. Annotation resources within DMRichR include the annotatr and rGREAT (Genomic Regions Enrichment of Annotations Tool) open source packages in Bioconductor. Annotatr (Cavalcante & Sartor, 2017) was used to visualize and compare annotated genomic sites/regions (e.g., promoters, 5’UTRs, exons, introns) that were identified within the discovery and validation sets, while rGREAT (Gu, 2014) was used for mapping genes to the sites. The mapped genes were then utilized for pathway and functional analyses using Ingenuity Pathway Analysis software (Qiagen, Germantown, MD) as described below.

### Gene ontology and network prediction analysis

Gene ontology analysis of the DMRs assigned to genes was accomplished using two different programs; GofuncR (Grote, 2020), which was modified for use with DMRichR, and the open-access STRING Version 11.0 (Search Tool for the Retrieval of Interacting Genes/Proteins) (Szklarczyk et al., 2019). GofuncR maps a DMR to a gene if it is between 5 kb upstream and 1 kb downstream of the gene body and also uses information from the background regions. A custom meta p-value analysis was performed using the sum of logs of the p-values of GO terms to integrate the data from the discovery and validation sets, and then terms with a meta p-value < 0.05 were slimmed using REVIGO (Supek, Bošnjak, Škunca, & Šmuc, 2011) to reduce redundancy among the most significant GO terms. Ingenuity Pathway Analysis (IPA) Version 01-13 (Qiagen, Germantown, MD) was used to discover pathways and functions enriched among the DMR-associated genes based on Fisher Exact p-values of ≤ 0.05, using the curated genes in IPA’s Knowledgebase as the reference gene set. The overall workflow for this study is summarized in **Figure 1**.

**Figure 1.**
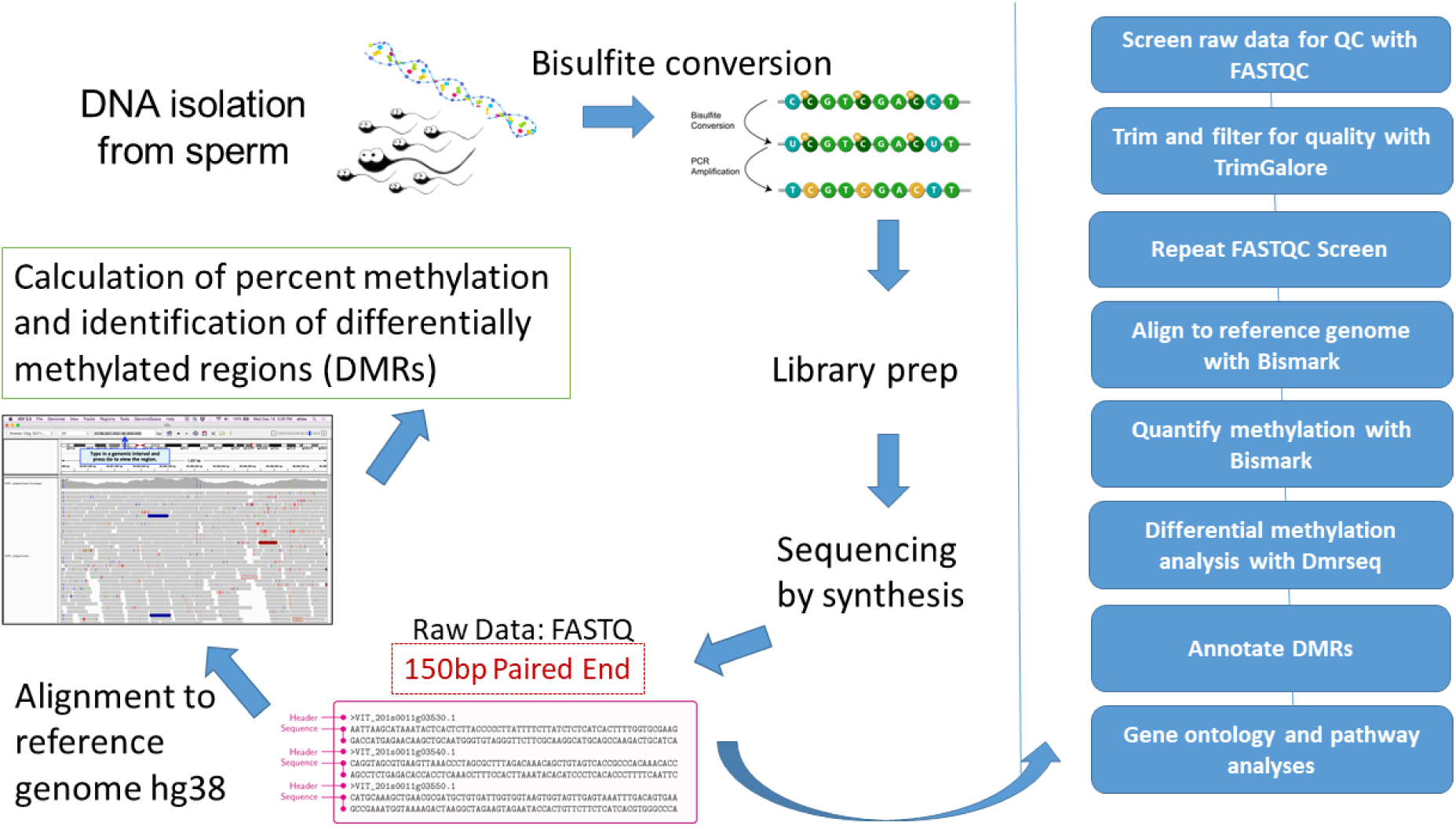
Overview of workflow for this study

### Class prediction analysis of WGBS data using machine-learning approaches

In order to select reliable predictors of DDE exposure levels for further validation by pyrosequencing, random forest (RF) and linear support vector machine (SVM) models were used to create binary classifiers that identify the exposure class (either ‘First’ or ‘Third’ tertile) of a given sample. A total of 261 DMRs from the analysis of all 52 samples, without ComBat batch correction, were utilized as predictors for the models. The samples were split according to sequencing batch, where the 32 samples from the first batch were used as a training set, and the 20 samples from the second batch were used as a testing set. The training set consisted of 16 samples in the ‘Third’ tertile and 16 matched controls labeled as the ‘First’ tertile. Similarly, the testing set consisted of 10 samples in each of these two tertile groups. Then, the machine learning model, either RF or SVM, was built with 5-fold cross-validation on the training set and used to predict the DDE exposure ‘Tertile’ class in the testing set.

To identify the most predictive DMRs for DDE exposure, we used two wrapper feature selection algorithms from the sigFeature Bioconductor package and the Boruta R package. Among the three types of feature selection methods (filter, embedded, and wrapper), we chose the wrapper method as it usually provides the most relevant feature set for a specific model. Wrapper methods rank features by repeatedly generating a subset of features from the full set of features and training a particular type of model using the generated subset.

There are many variable selection methods for the RF algorithm, but the Boruta algorithm was shown to be the most relevant, as it identifies all relevant variables, including variables that contain redundant information. In genomics studies, the inclusion of correlated and thus redundant variables may be important for model performance and the consistency of results(Degenhardt, Seifert, & Szymczak, 2019). The Boruta algorithm in the Boruta package applies the RF algorithm from the randomForest R package (Kursa & Rudnicki, 2010).

Of the variable selection methods that use the SVM algorithm, we decided to use the SVM recursive feature elimination (SVM-RFE) algorithm and t-statistic from the sigFeature package. SVM-RFE, one of the most effective feature selection methods, uses a greedy algorithm to find the best subset of features for binary classification tasks but does not consider differentially significant features between classes. The sigFeature package addresses this limitation by using the t-statistic to also find differentially significant features (Das, Roychowdhury, Das, Roychoudhury, & Tripathy, 2020).

### Pyrosequencing analyses

Pyrosequencing analyses of selected DMRs were performed in the Weksberg laboratory at The SickKids Research Institute of The Hospital for Sick Children; Toronto, Ontario, Canada. These DMRs included specific regions for *CSMD1, NRXN2, RBFOX1, MIRLET7BHG, PTPRN2, SNORD115-30,* and *SNORD115-37* that were identified by the WGBS analyses. Primers for each of the regions that typically included multiple methylation sites as well as the PCR conditions for the pyrosequencing analyses are provided in **Supplemental File 1**. The resulting pyrosequencing data were returned to us for further analyses of the methylation profiles of each region as a function of DDE serum concentration (µg/gm lipid) for each sample.

### Hypergeometric distribution analyses

Hypergeometric distribution analyses were employed to identify the significance of overlap between DMR-associated genes and autism risk genes from the SFARI Gene database (Abrahams et al., 2013). First, the overlapping genes were identified using a Venn diagram software program called Venny 2.1.0 https://bioinfogp.cng.csic.es/tools/venny/ (Oliveros, 2007). Significant overlap between the DMR-associated genes and SFARI genes was determined by hypergeometric distribution analyses using the CASIO Keisan Online Calculator <http://keisan.casio.com/exec/system/1180573201>, with significance determined by an upper cumulative Q-value of ≤ 0.05. These two programs were also used to identify significant overlap of DMR-associated genes from this study and those from other studies.

## Results

### DMRs associated with high and low exposures to DDE

WGBS analyses of the initial 32 samples (discovery set) with correction for all covariates revealed a total of 894 differentially methylated regions (DMRs, permutation p ≤ 0.05) between the first and third DDE exposure tertiles, while subsequent analyses of the validation set of 20 samples resulted in a total of 865 DMRs (**Figure 2**, DMRs in **Supplemental Tables 2 and 3)**. The overall distribution profiles of annotated gene and CpG regions were similar for both analyses (**Suppl. Figure 4**). Analysis of larger blocks of sequence for differential methylation revealed a single region that reached genome-wide significance after multiple testing correction (q < 0.022) when all 52 samples were combined (**Figure 3**). This block covered a 40,618 bp region of the *SNORD115* locus that is maternally imprinted, meaning expressed exclusively from the paternal allele. The validation data set shows nominal significance for increased methylation across this locus with a width of 63,606 bp (p < 0.011).

**Figure 2.**
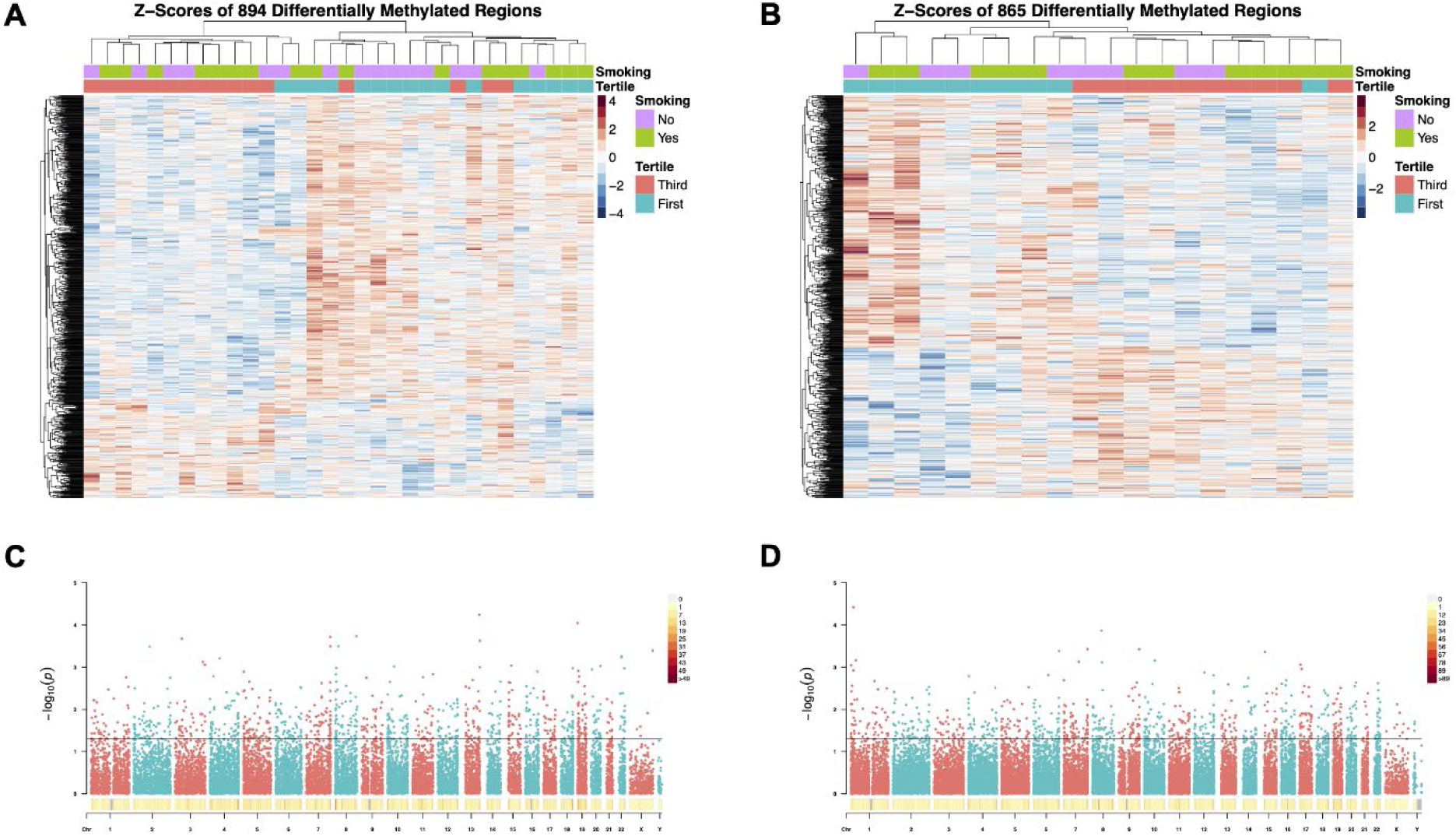
Heatmaps and Manhattan plots depicting the results of the WGBS analyses on the discovery set (A, C) and the validation set (B, D)

**Figure 3.**
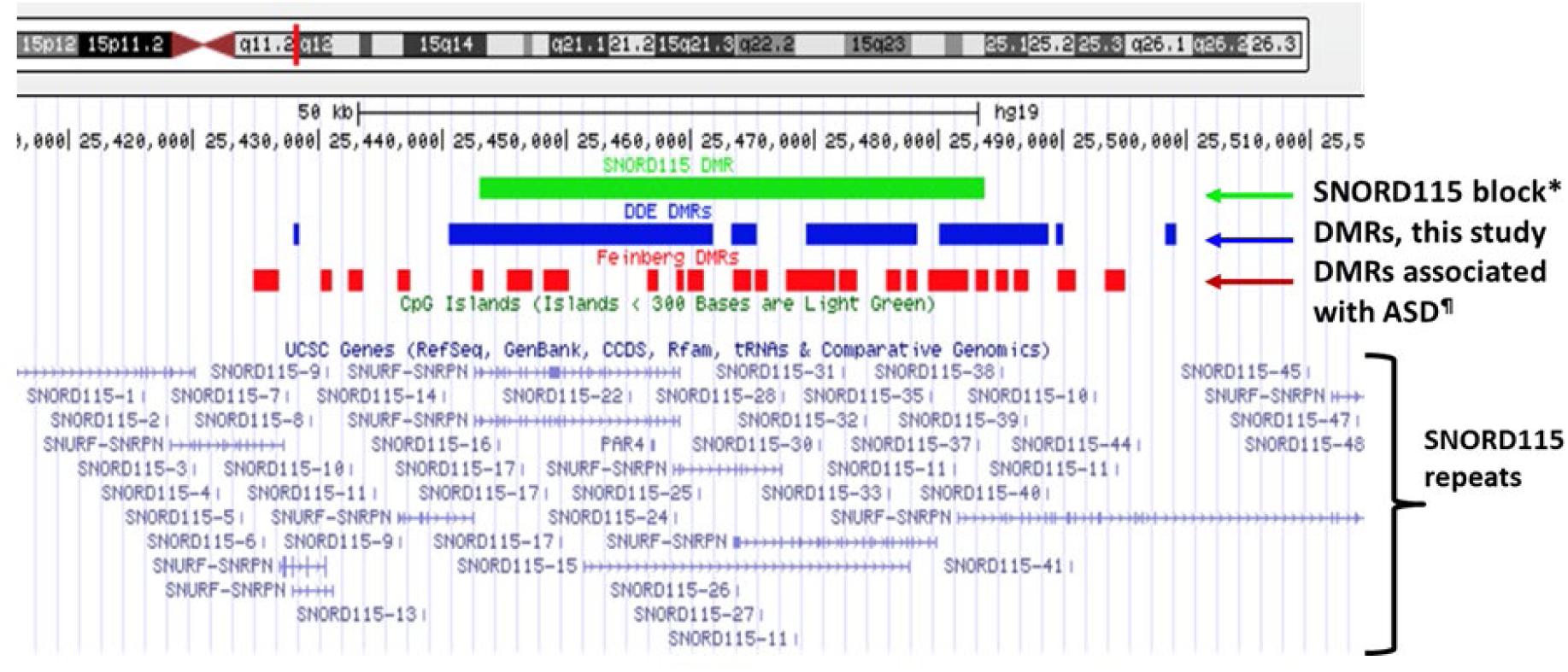
*SNORD115* region showing genome-wide significance for differential methylation after correction for multiple testing

### Sperm DMRs associated with neurodevelopmental processes and ASD-risk genes

Gene ontology analyses of the genes within the DMRs in both discovery and validation samples were performed using two different gene mapping and ontology approaches. GOfuncR analysis provides an overview of the top biological processes, cellular components, and molecular functions associated with the DMR-associated genes from both discovery and validation sets (**Figure 4**), while STRING analysis of these genes not only replicates some of the GO terms from the GOfuncR analysis but also reveals significant over-representation of a number of processes involved in nervous system development and function that are shared by both datasets (**Table 1**). Notably, these shared processes include nervous system development, generation of neurons, neurogenesis, neuron differentiation, and synapse organization. The enrichment in neuronal processes was further confirmed by pathway and functional analyses of the DMR-associated genes using IPA. CREB signaling in neurons, the endocannabinoid developing neuron pathway, netrin signaling, calcium signaling, GABA signaling, and axon guidance signaling are among the canonical pathways significantly enriched in both data sets (**Table 2**), while recognition of neurons, outgrowth of neurites, and neurotransmission are shared functions over-represented among the DMR-associated genes (**Table 3**).

**Figure 4.**
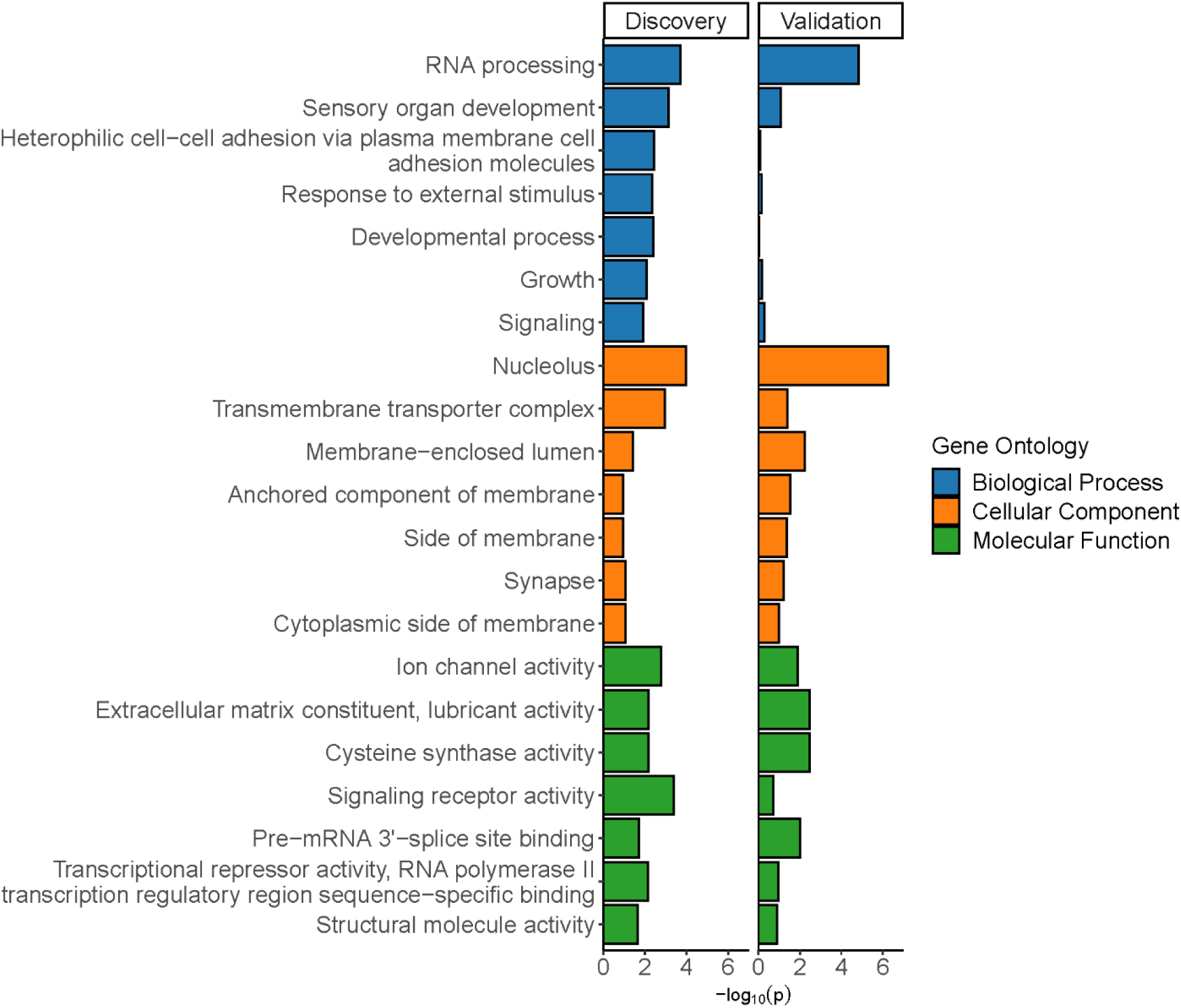
Bar plot of discovery and validation cohort specific p-values from a meta p-value analysis of the least dispensable significant (*p* < 0.05) gene ontology terms from GOfuncR

**Table 1.** Gene ontology terms enriched among DDE DMR-associated genes from discovery and validation sets.

| GO ID | Pathway description | Discovery FDR | Validation FDR |
| --- | --- | --- | --- |
| GO.0007399 | nervous system development | 1.80E-11 | 4.80E-03 |
| GO.0007275 | multicellular organism development | 2.13E-08 | 1.60E-03 |
| GO.0048731 | system development | 3.14E-08 | 4.10E-03 |
| GO.0048856 | anatomical structure development | 7.91E-08 | 2.20E-03 |
| GO.0022008 | neurogenesis | 5.51E-07 | 2.19E-02 |
| GO.0032502 | developmental process | 5.51E-07 | 5.50E-04 |
| GO.0048699 | generation of neurons | 5.51E-07 | 1.17E-02 |
| GO.0048666 | neuron development | 1.55E-06 | 4.12E-02 |
| GO.0030182 | neuron differentiation | 2.95E-06 | 2.19E-02 |
| GO.0000904 | cell morphogenesis involved in differentiation | 1.03E-05 | 3.89E-02 |
| GO.0048468 | cell development | 2.77E-05 | 1.48E-02 |
| GO.0030154 | cell differentiation | 1.50E-04 | 2.19E-02 |
| GO.0048869 | cellular developmental process | 2.50E-04 | 1.45E-02 |
| GO.0048513 | animal organ development | 2.80E-04 | 1.92E-02 |
| GO.0003279 | cardiac septum development | 7.10E-04 | 3.89E-02 |
| GO.0050808 | synapse organization | 1.80E-03 | 4.18E-02 |
| GO.0072359 | circulatory system development | 1.80E-03 | 4.12E-02 |
| GO.0007155 | cell adhesion | 2.40E-03 | 9.60E-03 |
| GO.0003148 | outflow tract septum morphogenesis | 3.30E-03 | 2.74E-02 |
| GO.0050793 | regulation of developmental process | 4.10E-03 | 2.60E-02 |
| GO.0050794 | regulation of cellular process | 5.80E-03 | 1.90E-04 |
| GO.0000122 | negative regulation of transcription by RNA polymerase II | 6.10E-03 | 7.70E-04 |
| GO.2000026 | regulation of multicellular organismal development | 6.10E-03 | 3.89E-02 |
| GO.0050789 | regulation of biological process | 1.04E-02 | 3.30E-04 |
| GO.0006928 | movement of cell or subcellular component | 1.50E-02 | 2.19E-02 |
| GO.0007423 | sensory organ development | 1.50E-02 | 1.05E-02 |
| GO.0048518 | positive regulation of biological process | 3.04E-02 | 3.80E-04 |
| GO.0050767 | regulation of neurogenesis | 4.36E-02 | 4.12E-02 |
| GO.0051960 | regulation of nervous system development | 4.54E-02 | 3.51E-02 |

**Table 2.**
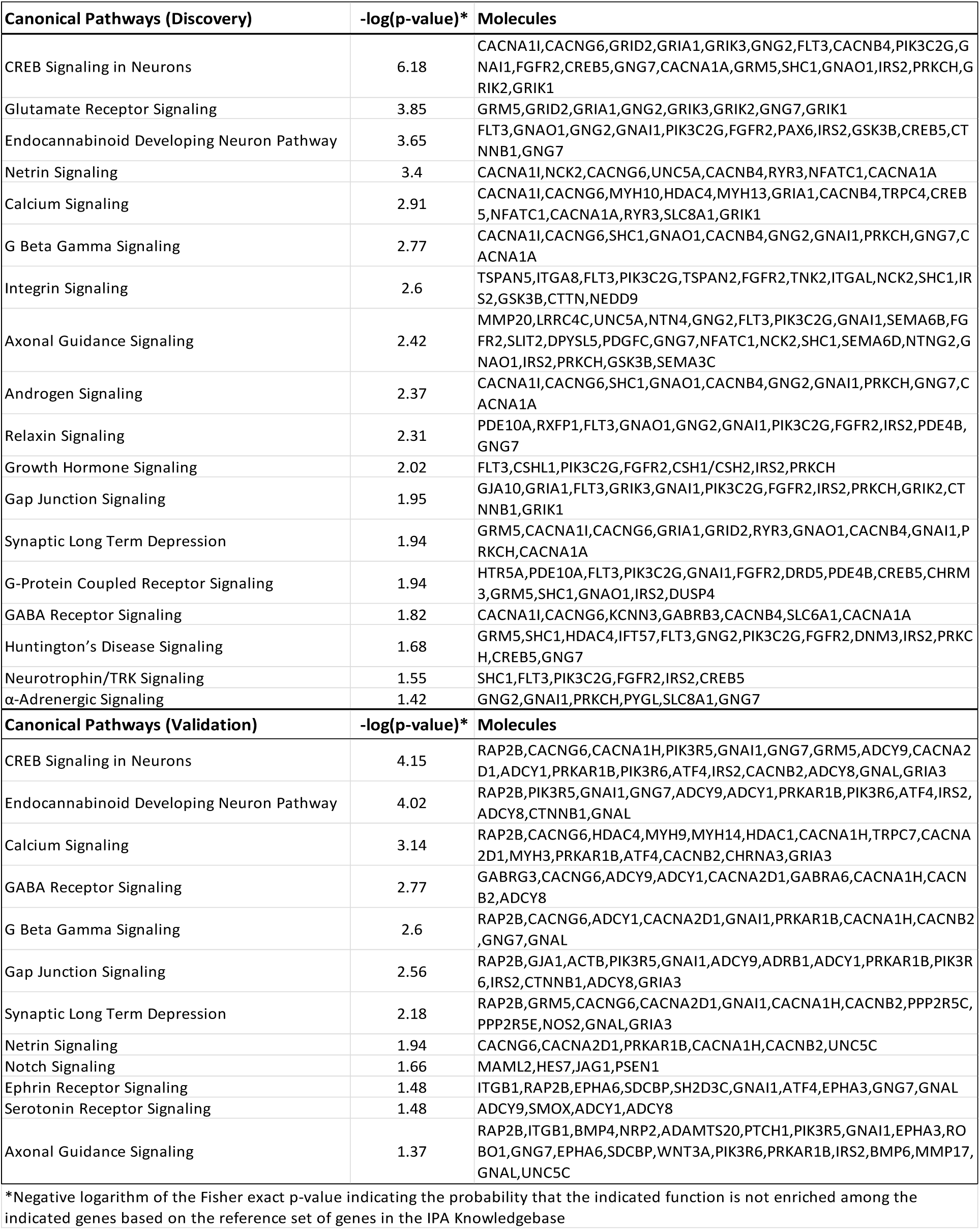
Canonical pathways enriched among DDE DMR-associated genes from discovery and validation sets.

**Table 3.**
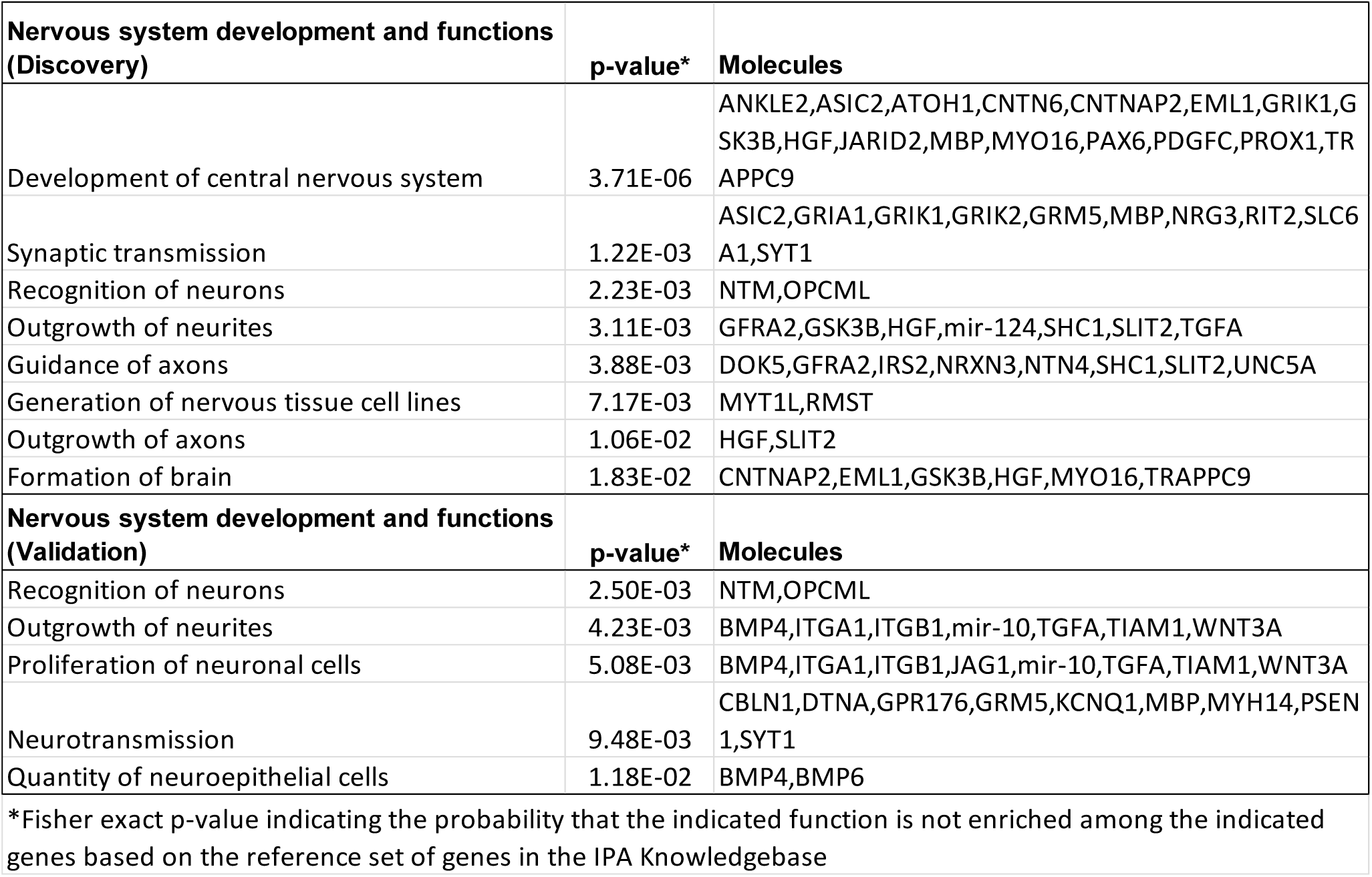
Nervous system functions enriched among DDE DMR-associated genes from discovery and validation sets.

| **Nervous system development and functions (Discovery)** | **p-value\*** | **Molecules** |
| --- | --- | --- |
| Development of central nervous system | 3.71E-06 | ANKLE2,ASIC2,ATOH1,CNTN6,CNTNAP2,EML1,GRIK1,GSK3B,HGF,JARID2,MBP,MYO16,PAX6,PDGFC,PROX1,TRAPPC9 |
| Synaptic transmission | 1.22E-03 | ASIC2,GRIA1,GRIK1,GRIK2,GRM5,MBP,NRG3,RIT2,SLC6A1,SYT1 |
| Recognition of neurons | 2.23E-03 | NTM,OPCML |
| Outgrowth of neurites | 3.11E-03 | GFRA2,GSK3B,HGF,mir-124,SHC1,SLIT2,TGFA |
| Guidance of axons | 3.88E-03 | DOK5,GFRA2,IRS2,NRXN3,NTN4,SHC1,SLIT2,UNC5A |
| Generation of nervous tissue cell lines | 7.17E-03 | MYT1L,RMST |
| Outgrowth of axons | 1.06E-02 | HGF,SLIT2 |
| Formation of brain | 1.83E-02 | CNTNAP2,EML1,GSK3B,HGF,MYO16,TRAPPC9 |
| **Nervous system development and functions (Validation)** | **p-value\*** | **Molecules** |
| Recognition of neurons | 2.50E-03 | NTM,OPCML |
| Outgrowth of neurites | 4.23E-03 | BMP4,ITGA1,ITGB1,mir-10,TGFA,TIAM1,WNT3A |
| Proliferation of neuronal cells | 5.08E-03 | BMP4,ITGA1,ITGB1,JAG1,mir-10,TGFA,TIAM1,WNT3A |
| Neurotransmission | 9.48E-03 | CBLN1,DTNA,GPR176,GRM5,KCNQ1,MBP,MYH14,PSEN1,SYT1 |
| Quantity of neuroepithelial cells | 1.18E-02 | BMP4,BMP6 |
| *Fisher exact p-value indicating the probability that the indicated function is not enriched among the indicated genes based on the reference set of genes in the IPA Knowledgebase |  |  |

To further investigate the relevance of these DMR-associated genes to ASD, we performed hypergeometric distribution analyses to determine enrichment in autism risk genes from the SFARI Gene database. **Figure 5** shows the overlap between DMR-associated genes from the discovery and validation analyses as well as the overlap between these DMR associated genes and the SFARI genes. The upper cumulative Q-values for enrichment in SFARI genes were 2.5 x 10^-7^ and 1.1 x 10^-5^ for the discovery and validation sets, respectively. Of the 138 overlapping DMR-associated genes between the discovery and validation sets, 14 are included in the SFARI Gene database. Notably, all of these genes are involved in development.

**Figure 5.**
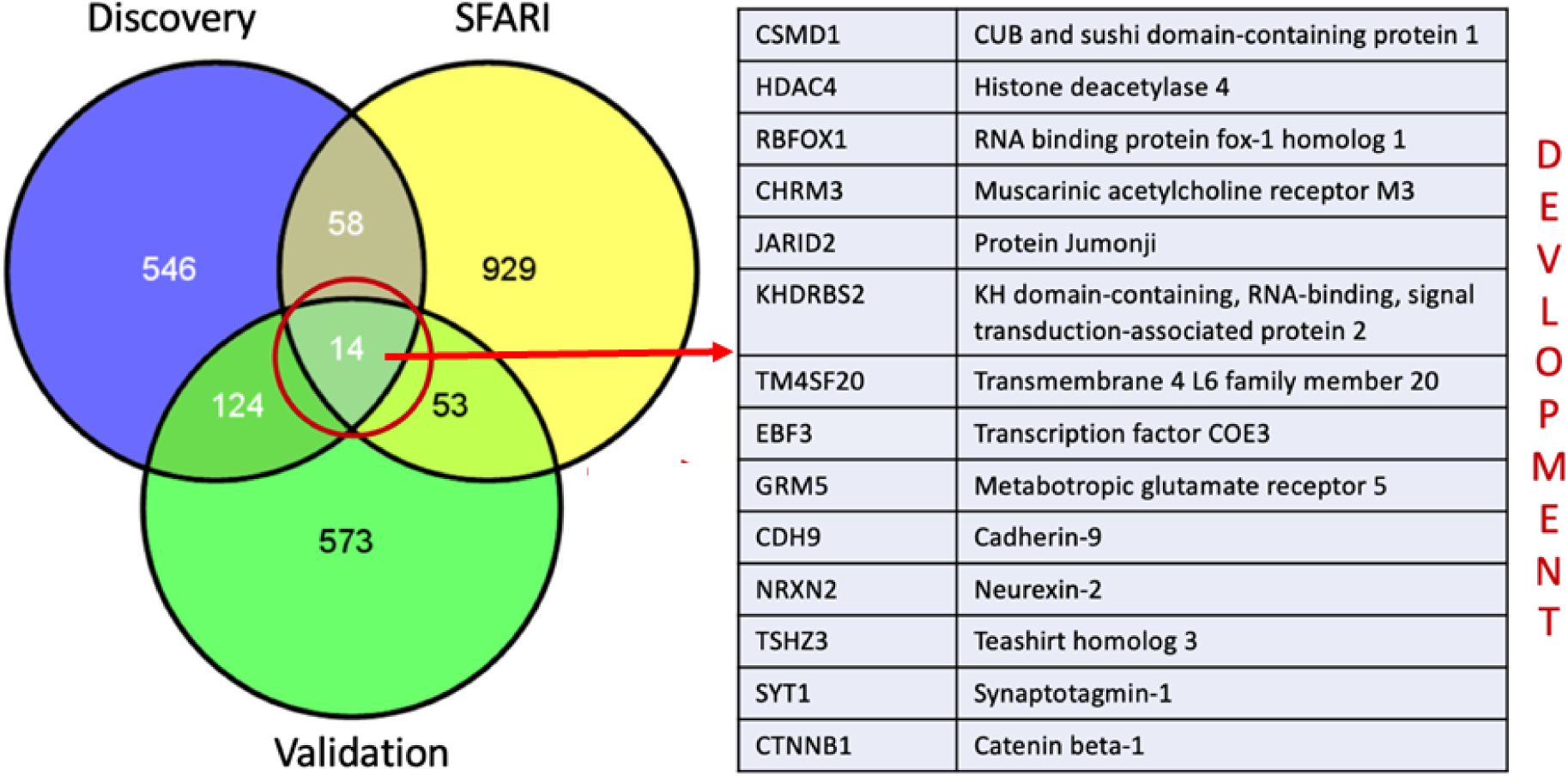
Venn diagram showing the overlap of DMR-associated genes among the discovery set, the validation set, and SFARI genes

### Class prediction analysis using a machine-learning approach

An attempt was made to identify DMRs that could reliably assign samples to the high or low exposure levels. Both random forest and linear support vector machine models were independently applied to the entire set of 261 DMRs obtained using all 52 samples (**Supplemental Table 4**). The discovery set of samples was used for training the classifier, and the validation set was used for testing the classifier. **Table 4** shows 15 of the top predictors resulting from these two classification approaches.

**Table 4.** List of top classifier genes for DDE tertile prediction from a combination of two separate machine learning analyses.

| Chr | start | end | annotation | distance to TSS | Gene Symbol | Gene Name |
| --- | --- | --- | --- | --- | --- | --- |
| chr2 | 98280512 | 98281225 | Intron 23 of 28 | 193396 | VWA3B | von Willebrand factor A domain containing 3B |
| chr2 | 240961395 | 240961697 | Intron 20 of 31 | 5754 | LOC200772 | uncharacterized LOC200772 |
| chr3 | 17133705 | 17133973 | Distal Intergenic | 123211 | PLCL2 | phospholipase C like 2 |
| chr3 | 43484849 | 43485664 | Intron 11 of 12 | 136404 | ANO10 | anoctamin 10 |
| chr4 | 2161268 | 2161626 | Intron 11 of 23 | 67605 | POLN | DNA polymerase nu |
| chr5 | 7927007 | 7928271 | Distal Intergenic | 30570 | MTRR | 5-methyltetrahydrofolate-homocysteine methyltransferase reductase |
| chr9 | 65389175 | 65390088 | Distal Intergenic | -103966 | FOXD4L5 | forkhead box D4 like 5 |
| chr10 | 14980617 | 14981682 | Intron 1 of 2 | 8062 |  | ?? |
| chr10 | 128066899 | 128068111 | Intron 11 of 20 | 159838 | PTPRE | protein tyrosine phosphatase, receptor type E |
| chr10 | 14989759 | 14992906 | Distal Intergenic | 17204 |  | ?? |
| chr13 | 110824201 | 110825915 | Distal Intergenic | -11117 | LINC00567 | long intergenic non-protein coding RNA 567 |
| chr13 | 114042350 | 114043198 | Intron 3 of 23 | 89413 | RASA3 | RAS p21 protein activator 3 |
| chr20 | 63948311 | 63949065 | Exon 2 of 3 | 7350 | UCKL1 | uridine-cytidine kinase 1 like 1 |
| chr22 | 46103014 | 46103671 | Exon 4 of 5 | 49145 | MIRLET7BHG | MIRLET7B host gene |
| chr22 | 46104978 | 46105643 | Exon 5 of 5 | 51109 | MIRLET7BHG | MIRLET7B host gene |

### Pyrosequencing analyses of several DMR-associated ASD risk genes

Several DMRs were selected for pyrosequencing validation. These included the regions harboring known autism risk genes, *CSMD1*, *NRXN2*, and *RBFOX1*, all of which were found to be differentially methylated in both discovery and validation analyses. In addition, we included *PTPRN2* which, like *NRXN2*, was found to be differentially methylated in cord blood from the Faroese population as a function of level of DDE exposure (Leung et al., 2018). Also selected for pyrosequencing were *MIRLET7HBG* which was implicated by the classifier analyses and two SNORDs (*SNORD115-30* and *SNORD115-37*) which are located in an imprinted region identified as differentially methylated in this study as well as in a previous study on paternal sperm from men with an autistic child (Feinberg et al., 2015). **Figure 6** shows the correlation curves for methyation level versus serum DDE exposure level for *CSMD1*, *NRXN2*, and *RBFOX1*. All of these genes showed a modest but statistically significant inverse correlation between methylation and DDE exposure levels, whereas *MIRLET7HBG* showed a trend towards increased methylation with increasing DDE levels (**Figure 7**). Similarly, consistent but not significant increases in methylation at multiple CpG sites within *SNORD115* were observed in the samples with higher exposures (**Table 5**). *PTPRN2*, on the other hand, showed no correlation between methylation detected by pyrosequencing and exposure levels.

**Figure 6.**
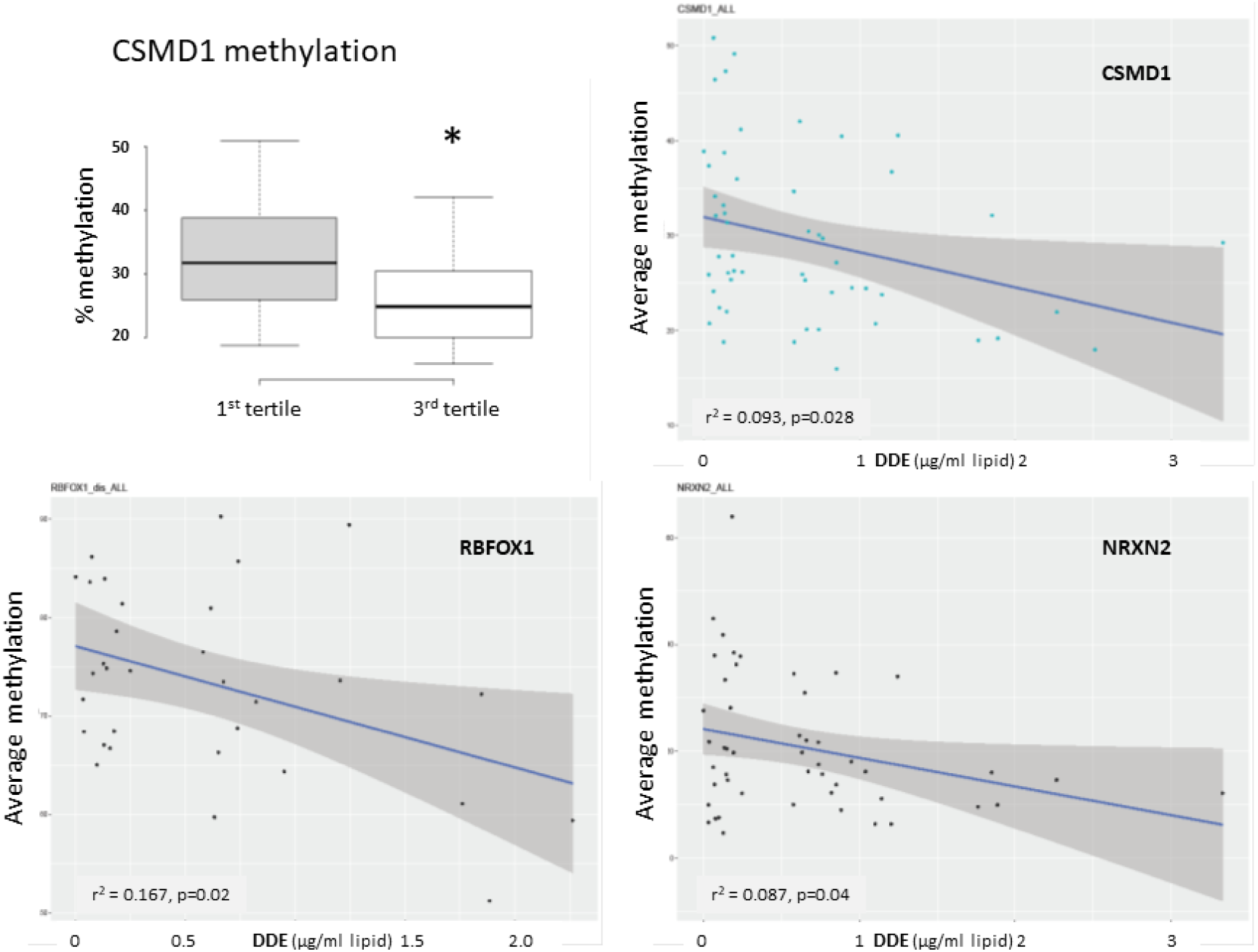
Results of pyrosequencing analyses of DMRs associated with *CSMD1* (A, B), *RBFOX1* (C), and *NRXN2* (D). The box plot (A) shows differential methylation at a single CpG site in *CSMD1* while the graphs show the average methylation as a function of DDE serum concentration (μg/gm lipid) for *CSMD1*, *RBFOX1* (10 sites, discovery set only), and *NRXN2* (7 sites, all samples in both discovery and validation sets). R-squared (r^2^) and p-values for the correlation curves are shown.

**Figure 7.**
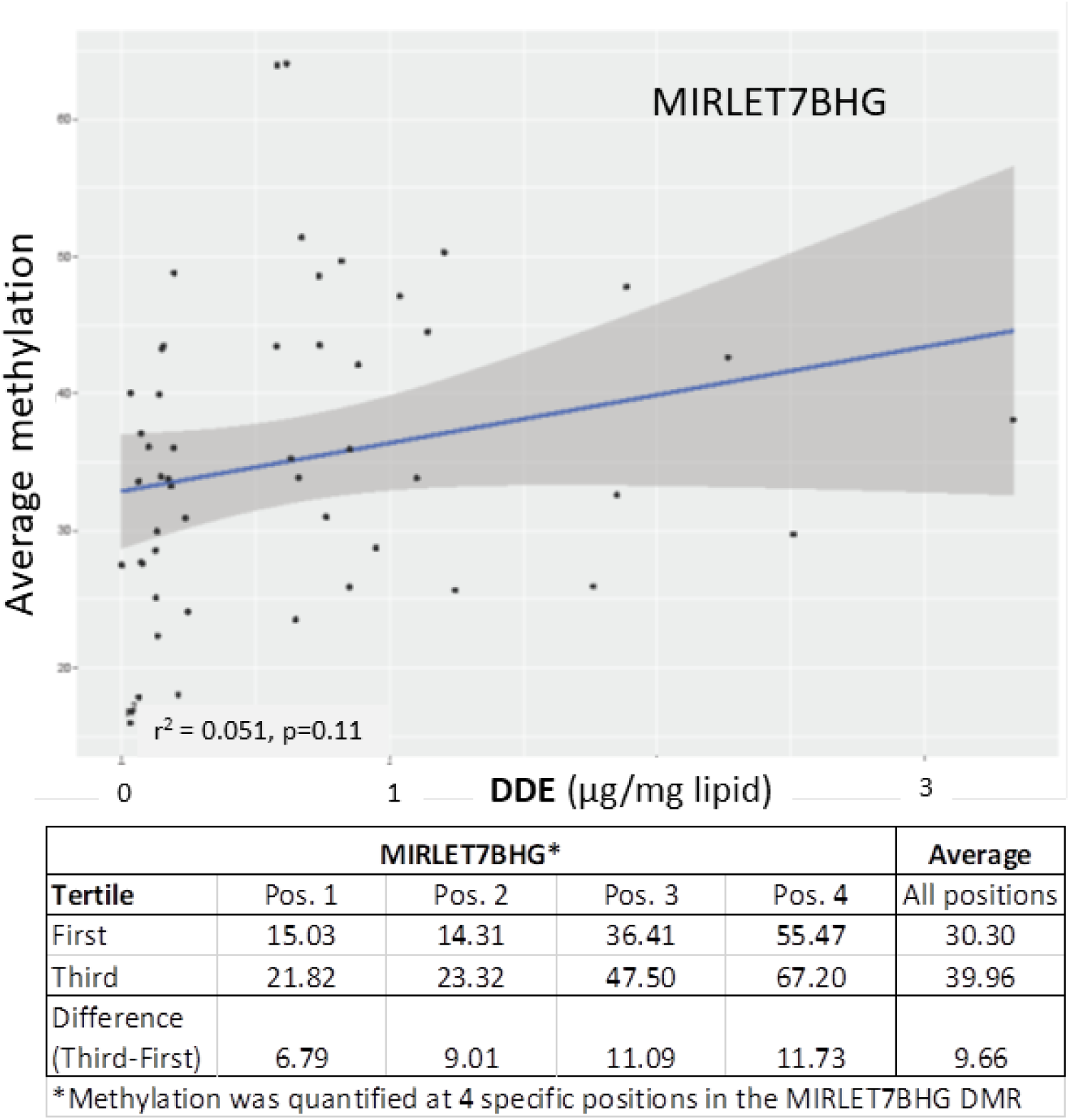
Results of pyrosequencing analyses of DMRs associated with *MIRLET7BHG.* The graph shows the average methylation across 4 CpG sites as a function of DDE serum concentration (μg/gm lipid) while the table below shows the methylation differences at each site.

**Table 5.**
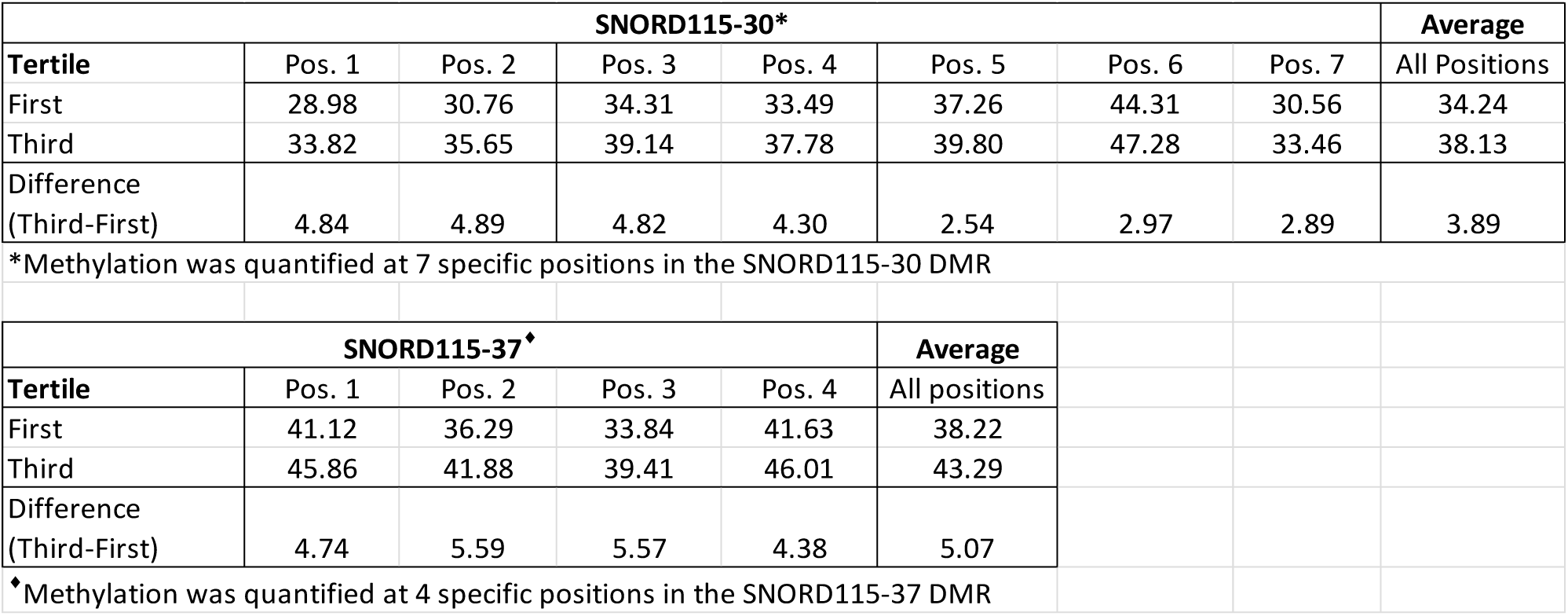
Differential methylation of *SNORD115-30* and *SNORD115-37* by DDE tertile validated by pyrosequencing analyses.

### Comparison of DMR-associated genes from this study with those from ASD-related and unrelated methylation studies

The DDE DMR-associated genes in sperm were compared to differentially methylated genes in a variety of tissues from studies on ASD-associated methylation differences. These tissues included cord blood from newborns, a fraction of whom was later diagnosed with ASD (Mordaunt et al., 2020), sperm (Feinberg et al., 2015) and placenta (Zhu et al., 2019) from parents of children with high risk for ASD, brain tissues from individuals with Dup15q syndrome that is often associated with ASD (Dunaway et al., 2016) as well as lymphoblastoid cell lines derived from individuals with a severe form of ASD (Hu, Hong, Xu, & Shu, 2020). Hypergeometric distribution analyses show that the DMR-associated genes from both the discovery and validation WGBS analyses overlapped significantly with those from ASD cord blood, paternal sperm, placenta, and Dup15q brain that also showed detectable PCB 95 exposures (Dunaway et al., 2016) (**Table 6**). DMR-associated genes from the discovery, but not validation, set also overlapped significantly with those from lymphoblastoid cell lines. Interestingly, the significance of the overlap between the DDE DMR-associated genes is highest in DMRs identified from cord blood from children diagnosed with ASD and lower in the more differentiated or immortalized tissues, i.e., brain and lymphoblasts, respectively. In the study on placental methylation, placentas of high-risk mothers with a child (or children) already diagnosed with ASD were obtained after the birth of a subsequent child for WGBS analysis (Schroeder et al., 2016). Subsequent analysis of DMRs from the placentas for ASD outcome revealed 596 nominally significant genes, of which two (*CYP2E1* and *IRS2*) reached genome-wide significance (Zhu et al., 2019). For the sperm methylation analysis, semen samples from fathers of a child already diagnosed with ASD, were obtained for methylation analyses (Feinberg et al., 2015). The placenta and sperm studies revealed DMR-associated genes related to ASD outcome or higher risk for ASD, respectively, in offspring of the high-risk parents in comparison to parents of neurotypical children, while the DMR-associated genes in cord blood may be more directly related to ASD diagnosis in the individual. In addition, the DDE DMR-associated genes from both discovery and validation datasets significantly overlapped with those from pan-cancer studies (Su et al., 2018). The overlap with cancer genes is not surprising since organochlorine exposures are well-known risk factors for cancer; thus, the current study reveals exposure-associated epigenetic changes in DMR-associated genes that may also increase cancer risk (Dorgan et al., 1999; Jaga & Dharmani, 2005; Purdue, Hoppin, Blair, Dosemeci, & Alavanja, 2007). Interestingly, cancer and ASD share many risk genes and pathways (Crawley, Heyer, & LaSalle, 2016), some of which may be influenced by environmental factors. On the other hand, there was no significant overlap of DDE DMR-related genes with those in cord blood from the Faroese population that were also associated with DDE exposures (Leung et al., 2018), although DMR-associated genes from both sperm (discovery set) and cord blood showed enrichment in multiple canonical pathways associated with neurological functions (**Table 7**). Comparison of the DMR-associated genes from the ASD and Faroese cord blood studies showed no significant overlap (Q = 0.84), but both sets of DMRs were highly enriched in genes on the X-chromosome. Intriguingly, the X-linked genes from the ASD cord blood were predominantly found in females (Mordaunt et al., 2020), while those from the Faroese cord blood were exclusively male specific (Leung et al., 2018). By comparison, despite the highly significant overlap between the DMR-associated genes in the Faroese sperm and the ASD cord blood, there were relatively few X-linked DMR-associated genes found in sperm. Collectively, these results suggest that exposure to persistent organic pollutants, such as DDE, is associated with altered methylation status of genes in sperm and early developmental tissues that are critically associated with ASD.

**Table 6.** Significant overlap among DMR-associated genes from different ASD studies and tissues compared to the DDE DMR-associated genes from this study.

| Samples for comparison of DDE DMRs (sperm)<br>(# DMR-associated genes) | Hypergeometric distribution Q-value |  |
| --- | --- | --- |
|  | Discovery (742) | Validation (763) |
| Cord blood from newborns later diagnosed with ASD<sup>1</sup> (2173) | 2.96E-23 | 8.56E-10 |
| Sperm (high ASD risk)<sup>2</sup> (144) | 1.22E-06 | 9.95E-05 |
| Placenta (ASD outcome)<sup>3</sup> (596) | 1.56E-04 | 6.78E-05 |
| Brain (Dup15q)<sup>4</sup> (942) | 1.90E-03 | 2.00E-03 |
| Lymphoblastoid cell lines<sup>5</sup> (827)* | 1.90E-02 | 8.00E-02 |
| Pan-Cancer<sup>6</sup> (434)<sup>♦</sup> | 7.34E-12 | 2.17E-09 |
| Faroese cord blood (DDE associated)<sup>7</sup> (1136) | 0.608 | 0.55 |
| <sup>1</sup> Mordaunt et al., 2020 |  |  |
| <sup>2</sup> Feinberg et al., 2015 |  |  |
| <sup>3</sup> Zhu et al., 2019 |  |  |
| <sup>4</sup> Dunaway et al., 2016 |  |  |
| <sup>5</sup> Hu et al., 2020; *males only, severely language-impaired |  |  |
| <sup>6</sup> Su et al., 2018; <sup>♦</sup>hypermethylated canyon genes |  |  |
| <sup>7</sup> Leung et al., 2018 |  |  |
| Note: Ref #s will be revised after final list of refs is generated |  |  |

**Note: the superscript ref #s will be changed to whatever ref # applies to the respective study in this manuscript.**

**Table 7.** Canonical pathways enriched among overlapping DDE DMR-associated genes between Faroese sperm and cord tissue.

| **Canonical Pathways** | **-log(p-value)\*** | **Molecules** |
| --- | --- | --- |
| Netrin Signaling | 3.53 | CACNA1I,NCK2,UNC5A |
| GABA Receptor Signaling | 3.06 | CACNA1I,GABRB3,KCNN3 |
| Circadian Rhythm Signaling | 2.71 | CREB5,VIP |
| CREB Signaling in Neurons | 2.02 | CACNA1I,CREB5,GRIK2 |
| Axonal Guidance Signaling | 1.82 | NCK2,NTN4,NTNG2,UNC5A |
| Cellular Effects of Sildenafil (Viagra) | 1.58 | KCNN3,PDE4B |
| GP6 Signaling Pathway | 1.55 | COL25A1,FGA |
| Corticotropin Releasing Hormone Signaling | 1.51 | CACNA1I,CREB5 |
| Extrinsic Prothrombin Activation Pathway | 1.5 | FGA |
| Gustation Pathway | 1.44 | CACNA1I,PDE4B |
| Methionine Degradation I (to Homocysteine) | 1.41 | MGMT |
| Protein Kinase A Signaling | 1.39 | CREB5,PDE4B,PTPRS |
| Cysteine Biosynthesis III (mammalia) | 1.37 | MGMT |
| GNRH Signaling | 1.35 | CACNA1I,CREB5 |
| Ephrin Receptor Signaling | 1.31 | CREB5,NCK2 |
| *Negative logarithm of the Fisher exact p-value indicating the probability that the indicated pathway is not enriched among the indicated genes based on the reference set of genes in the IPA Knowledgebase |  |  |

## Discussion

### Methylation patterns of sperm DNA are associated with DDE exposure levels

This study shows that the DNA methylation status in sperm may be influenced by lifelong exposure to environmentally derived persistent EDCs, such as DDE. The Faroese cohort used in this study is particularly exposed to higher than average levels of EDCs as a result of their natural diet which includes substantial amounts of pilot whale meat and blubber. Fatty tissues are reservoirs for lipophilic molecules, which include a wide variety of organochlorines, such as DDE as well as PCBs. The high correlation between the levels of multiple organochlorines and DDE in serum indicates that the DDE exposures employed in this study are proxies for exposures to persistent organochlorines in general. Although some of these compounds have now been banned for use, the long half-lives of such compounds and/or their breakdown products still pose risk of environmental exposures.

### DMRs harbor genes enriched for neurodevelopment and function

Although there are hundreds of genes whose methylation is altered by elevated exposure to DDE and other organochlorines, our gene ontology and pathway analyses reveal that genes involved in nervous system development and function are among the most significantly over-represented in DMRs. Moreover, a significant number of these genes are also autism risk genes that are included in the SFARI Gene database. Among the ASD-risk DMR-associated genes validated by pyrosequencing are *CSMD1*, *NRXN2*, and *RBFOX1*. *CSMD1*, which encodes for CUB and Sushi multiple domains 1, is highly expressed in brain tissues where it has been associated with neuronal growth cone stabilization and neuritogenesis (Molenaar et al., 2012). Aside from being a risk gene for ASD (Cukier et al., 2014; Guo et al., 2017; Hu, Addington, & Hyman, 2011), it has also been implicated in schizophrenia, bipolar disorder, and post-traumatic stress disorder (Woo, Yu, Kumar, & Reifman, 2017). *NRXN2* codes for neurexin 2, a brain-enriched cell adhesion molecule that has long been associated with ASD (Dachtler et al., 2015; Gauthier et al., 2011; Mohrmann, Gillessen-Kaesbach, Siebert, Caliebe, & Hellenbroich, 2011). NRXN2 plays a role in early cortical synaptogenesis and axon guidance (Harkin et al., 2017). *RBFOX1* encodes for RNA binding fox-1 homolog 1, a neuron-specific splicing factor that has been implicated in many studies on ASD (Bacchelli et al., 2020; Griswold et al., 2015; Martin et al., 2007). Interestingly, *RBFOX1* was one of the top transcriptional targets of the orphan nuclear hormone receptor RORA which we found to be regulated by sex hormones (Sarachana & Hu, 2013) as well as EDCs, including DDE and atrazine, an herbicide (Shu, Kocher, and Hu, unpublished data).

### EDC-associated differential methylation also impacts noncoding regions of the genome

*MIRLET7BHG* codes for a long noncoding RNA (lncRNA) whose neonatal umbilical cord methylation level has been associated with birth weight in a study of the effects of prenatal environment and genotype on offspring weight and obesity in early childhood (Lin et al., 2017). This gene was identified in this study as a potential predictor of exposure level by machine-learning analyses. Interestingly, this gene was one of eight lncRNAs that were differentially expressed between a group of women with polycystic ovary syndrome (PCOS) and control women, and the only one whose expression was correlated with BMI (Butler et al., 2019). As PCOS is associated with abnormally high levels of male hormones, it is notable that the methylation status of *MIRLET7BHG* is influenced by EDC exposures.

The altered methylation in a large block on chromosome 15 encompassing the noncoding *SNORD115* region is of particular interest inasmuch as this region was also found to be differentially methylated in the sperm of fathers with a child exhibiting ASD in comparison to the sperm of fathers of unaffected children (Feinberg et al., 2015). However, the origins of such differences in sperm DNA methylation that are associated with ASD are unknown. The results from this study suggest that environmental exposures to certain EDCs may in part be responsible for alterations in the sperm methylome and, in particular, the *SNORD115* region. This region is imprinted maternally and normally shows compact heterochromatin until its expression, which is exclusive to neurons. Intriguingly, the *SNORD115* locus is not expressed in any tissue besides the brain. With respect to function, the *SNORD115* region on human chromosome 15q11-q13 contains a series of 48 highly conserved small nucleolar RNAs (snoRNAs) that participate in the modification of other noncoding RNAs and site-specific 2′-O-methylation of substrate RNAs (Galardi et al., 2002) as well as alternative splicing and RNA editing, especially of 5-HTR2C pre-mRNA (Bratkovič, Modic, Camargo Ortega, Drukker, & Rogelj, 2018; Cavaillé, 2017; Raabe et al., 2019). In addition, a previous study has also identified alterations at this locus in association with exposures to EDCs, albeit of a more transient (non-persistent) nature. Specifically, increased methylation was observed in the *SNORD115* locus in human fetal lung tissue of discontinued pregnancies of women exposed to BPA (Faulk et al., 2015).

Although deletions in this genomic region are associated with Prader-Willi syndrome (PWS), loss of *SNORD115* alone is not sufficient to cause the disease (Bürger, Horn, Tönnies, Neitzel, & Reis, 2002; Runte, Varon, Horn, Horsthemke, & Buiting, 2005). Maternal duplications in the chr15q11-q13 region (aka. Dup15q syndrome) have also been associated with ASD as well as other developmental disorders, with some genes showing altered methylation status (Depienne et al., 2009; Finucane et al., 2016; Scoles, Urraca, Chadwick, Reiter, & Lasalle, 2011). As mentioned earlier, Dup15q was shown to be a strong predictor of PCB 95 exposure (Mitchell et al., 2012). The Mitchell et al. study further showed that LINE-1 methylation was reduced in Dup 15q and PWS samples but not idiopathic ASD, suggesting gene x environment interactions possibly mediated through epigenetic modifications in the genetically defined but not idiopathic ASD. However, *SNORD115* genes were not specifically implicated in these studies. Thus, the present study, coupled with that of Feinberg et al. on sperm from fathers of a child with ASD (Feinberg et al., 2015), reveals an additional epigenetic mechanism through which long-lived EDCs may mediate ASD-related changes in this region. Moreover, such changes in germline cells raise the possibility of transgenerational inheritance of phenotype as described in multiple animal studies.

### Potential significance of methylation changes in sperm cells

Previous studies on animal models have reported transgenerational effects of EDC exposures on disease and behavioral phenotypes, some of which were shown to be mediated by epigenetic changes in the germline (Anway & Skinner, 2008; Manikkam et al., 2012; McBirney et al., 2017; Skinner, Manikkam, & Guerrero-Bosagna, 2011). Initial findings in rodents have shown that exposure to vinclozolin (an androgenic EDC) or methoxychlor (an estrogenic EDC) led to increased male infertility and related characteristics such as decreased sperm count in all subsequent generations, from F1 through F4 (Anway, Cupp, Uzumcu, & Skinner, 2005). In addition, when gestating F0 females were given various doses of a mix of EDCs during embryonic development, F1 and F3 generations exhibited increased total disease. Differentially methylated regions were found in sperm of the the F3 generation that included promoters of genes associated with underlying diseases such as obesity, PCOS, and ovarian disease (Manikkam, Tracey, Guerrero-Bosagna, & Skinner, 2013). These studies and others (McBirney et al., 2017) indicate that F0 exposure can lead to transgenerational effects on phenotypes that were associated with epigenetic changes in sperm that were specific to adult onset disease.

### Advantages and limitations of this study design and future directions

A primary advantage of this study on sperm from men with different levels of exposure to DDE is that the Faroe Islands population is considered a “genetic isolate”, thus reducing genetic heterogeneity that is often a major challenge in epigenetic studies. Another advantage is the natural Faroese diet, rich in pilot whale meat and blubber, that exposes the population to higher than average levels of persistent organochlorines. While we show differences in methylation patterns in sperm from men with high versus low exposures to DDE (which were highly correlated with the sum of the four most prevalent PCB congeners), an obvious limitation of this study is the lack of a completely unexposed set of samples for comparisons. Because EDCs are known to exhibit non-monotonic dose-response curves (Vandenberg et al., 2012), it is possible that the lowest exposures examined here may show even greater methylation differences relative to unexposed controls than the differences between the first and third exposure tertiles included in this study. Moreover, we were not able to correlate methylation changes with changes in gene expression in the same tissues inasmuch as spermatocytes are transcriptionally inactive. Additionally, we could not correlate EDC exposure with ASD risk in offspring, as there was no information on children (if any) of the men in this Faroese cohort. Such a study would be particularly interesting as it has been reported that neurobehavioral deficits in a Faroese birth cohort of 7-year-old children are associated with prenatal exposures to organochlorine neurotoxicants in seafood as measured in umbilical cord tissue (Grandjean et al., 2001). Aside from investigating the possible relationships between organochlorine exposures, sperm DNA methylation profiles, and neurodevelopmental disorders, the availability of this publicly accessible methylation data on DDE/organochlorine-associated changes in the sperm methylome of exposed individuals will nevertheless provide a valuable resource for studies on other diseases and conditions, such as cancer, obesity, diabetes, and infertility, which are also linked to environmental exposures.

Future studies should include additional cohorts sampled longitudinally to monitor temporal changes in individual sperm methylation levels as a function of cumulative EDC exposures as well as expanded concentration levels of DDE/organochlorines. Given the association between exposure levels of DDE and altered methylation of many neurodevelopmental genes, it will also be of interest to investigate the relationships between DDE exposures of the men, epigenetic changes in sperm, and the health outcomes of their children.

## Conclusions

This study shows that elevated exposure to DDE, one of a class of persistent organochlorines, is associated with differential genome-wide DNA methylation patterns in sperm when compared against the lowest exposure levels. The DMRs are enriched for genes involved in many biological processes, including neurological functions and pathways impacted by neurodevelopmental disorders. This study thus supports the link between environmental EDC exposures and epigenetic changes in germ cells that may impact the regulation of many genes associated with disease phenotypes, including ASD. Studies involving animal models have shown that DNA methylation patterns as well as associated phenotypes or diseases can be stably and heritably transmitted through the germline in several generations of offspring, specifically in relation to EDC exposure. It is unknown whether the DNA methylation differences noted in this study arose from direct exposure of the individual sperm donors to EDCs or from ancestral exposures that led to the inheritance of specific DNA methylation patterns in the donors’ sperm.

## Data Availability

All raw and processed data from whole genome bisulfite sequencing has been deposited into GEO (GEO accession # GSE165915).

## Acknowledgements

This study was supported by the National Institute of Environmental Health Sciences of the National Institutes of Health (grant R21 ES028124 to VWH and partial support from R01 ES029213 to JML). BIL was supported by a Canadian Institutes of Health Research (CIHR) postdoctoral fellowship (MFE-146824) and a CIHR Banting postdoctoral fellowship (BPF-162684). We thank Dr. Pál Weihe (Department of Occupational Medicine and Public Health, Faroese Hospital System, Tórshavn, Faroe Islands) for generously providing the semen samples, demographic information on the semen donors, and data on serum levels of DDE and other organochlorines that had been determined previously by Dr. Philippe Grandjean (Department of Environmental Medicine, University of Southern Denmark, Odense, Denmark). We also thank Dr. Melissa Perry (Department of Environmental and Occupational Health, GWU) for dividing the semen samples from the Faroese cohort into exposure tertiles based on DDE serum levels which allowed us to process the samples upon receipt from sperm isolation through bisulfite sequencing without knowledge of the donor’s exposure group.

## Authors’ contributions

AGM was responsible for sequence alignment, quality control, WGBS, and bioinformatics analyses of the DMRs. HTS was responsible for sample organization and preparation, including sperm and DNA isolation from the semen aliquots. BIL also contributed substantially to WGBS and bioinformatics analyses. HH performed the machine learning analysis. CB analyzed the pyrosequencing data. YL provided statistical support and advice. JML provided advice and discussion of the WGBS analysis and data interpretation. VWH conceived of the study, performed bioinformatics and hypergeometric analyses on the DMR-associated genes, and wrote the manuscript. AGM, BIL, and JML also contributed to manuscript preparation and editing.

## List of Supplemental Materials

### Supplemental Tables

Table S1. Summary of demographic and EDC data for sperm donors in each exposure tertile

Table S2. DMRs identified in the discovery set

Table S3. DMRs identified in the validation set

Table S4. DMRs identified in the combined set of 52 samples

### Supplemental Figures

Figure S1. Correlation between serum concentrations of DDE and DDT and between DDE and the sum of 4 PCBs (PCB118, 138, 153, and 180)

Figure S2. Relationship between DDE serum concentrations and BMI and smoking status

Figure S3. Correlation between DDE serum concentrations and sperm parameters

Figure S4. Annotated genomic sites/regions among DMRs in discovery and validation sets

**Supplemental File 1:** Primer sequences and PCR conditions for pyrosequencing analyses

